# Gender dependent pharmacotherapy for blocking transition to chronic back pain: a proof of concept randomized trial

**DOI:** 10.1101/19006627

**Authors:** Diane Reckziegel, Pascal Tétreault, Mariam Ghantous, Kenta Wakaizumi, Bogdan Petre, Lejian Huang, Rami Jabakhanji, Taha Abdullah, Etienne Vachon-Presseau, Sara Berger, Alexis Baria, James W Griffith, Marwan N Baliki, Thomas J Schnitzer, A Vania Apkarian

**Affiliations:** Center for Chronic Pain and Drug Abuse, Northwestern University Feinberg School of Medicine, Chicago, USA; Department of Physiology, Northwestern University Feinberg School of Medicine, Chicago, USA; Shirley Ryan AbilityLab, Chicago, USA; Department of Medical Social Sciences, Northwestern University Feinberg School of Medicine, Chicago, USA; Department of Rheumatology, Northwestern University, Feinberg School of Medicine, Chicago, USA; Department of Physical Medicine and Rehabilitation, Northwestern University Feinberg School of Medicine, Chicago, USA; Department of Anesthesia, Northwestern University Feinberg School of Medicine, Chicago, USA

## Abstract

Preventing transition to chronic back pain (CBP) is an ideal strategy that would rescue patients from years to a lifetime of suffering with pain. Recent studies suggest involvement of sexually-dimorphic dopaminergic-motivational circuits in the transition to chronic pain (tCBP), and hints the combination of carbidopa/levodopa and naproxen (LDP+NPX) may block tCBP. We tested these concepts in early onset BP, who were stratified by risk for tCBP using brain properties. Those identified as low-risk entered a no-treatment arm. The rest were randomized into a double-blind, placebo and naproxen (PLC+NPX) controlled trial of oral LDP+NPX for 12 weeks, and a post-treatment 12-weeks follow-up. 59 participants completed the study. Both treatments resulted in ∼50% pain relief for ∼75%, sustained post-treatment. LDP+NPX was highly effective in females (>80% pain relief), it modified BP personality, and was related to objective brain functional changes. Although performed in a small group of early onset BP, multiple subjective and objective measures consistently suggest that these long-duration treatments persistently, and gender-dependently, block tCBP.

---

Chronic pain dramatically diminishes the quality of everyday life in about 20% of the world population, and for 100 million American adults^1^. As a large proportion of these patients are treated with opioids, chronic pain remains a primary contributor to the U.S. opiate epidemic^2^. Once established, its reversal becomes very difficult. Available treatments for chronic pain do not cure the condition, and majority of patients remain dissatisfied with their treatments. Thus, there is universal consensus that new, non-opioid, and efficacious treatments, especially for back pain, are urgently needed. Chronic low back pain (CBP) is the most prevalent type of chronic pain, and its brain properties are now well characterized.^3-6^ The current trial was limited to subjects with back pain persistent over multiple weeks, that is, subacute back pain (SBP), as such patients have a high probability of developing CBP^3,4,7^ and their pain is within the time window where it may be more malleable and thus reversible with proper treatment. Treatments that could block transition to chronic pain are an ideal preventative strategy to combat back pain, as they spare such patients from years of disability, diminish probability of exposure to opiate treatments, and decrease the associated staggering healthcare cost (estimated to be >$500 billion just in the USA).

Mesocorticolimbic brain (MCL) properties are a dominant risk factor for the transition from SBP to CBP^3,4,8,9^, and derived models account for >60% of the variance for risk of transitioning to CBP. Complimentarily, in rodent models of chronic pain, components of this circuit shows a causal relationship with pain-like behaviors, specifically neuronal activity in the medial prefrontal cortex and nucleus accumbens.^10,11^ Within nucleus accumbens, peripheral chronic neuropathic injury is accompanied with decreased concentration of dopamine, changes in excitability of dopamine D2 receptor expressing spiny neurons, shrinkage of dendrites and decreased synaptic inputs,^11^ as well as diminished dopamine D1 and D2 receptor expression.^12^ When such animals are treated with a combination of levodopa/carbidopa and naproxen (LDP+NPX) for multiple days, pain-like behaviors are diminished and many of the abnormalities of nucleus accumbens D2 spiny neurons are reversed.^11^ Importantly, treatment with either compound separately were not effective.^13^ This combination treatment was highly effective within days after the peripheral injury, but diminished in efficacy at later times.^13^ Yet, no clinical trial has studied efficacy of this strategy in relieving acute or chronic pain. Therefore, we reasoned that the regiment may be a potential therapy for treating SBP patients, testing the concept that early aggressive (3-months) treatment of SBP can prevent transition to CBP. Furthermore, most chronic pain patients are women,^14-16^ and MCL shows multiple lines of evidence for sexual dimorphism.^17^ In particular, brain dopamine release is lower in females.^18^ Therefore, we also reasoned that efficacy of the combination pharmacotherapy in SBP would be gender dependent, as we presumed the treatment being tested engages dopaminergic MCL circuits.

Thus, the primary aim of this study (NCT01951105) was to test the efficacy and toxicity of LDP+NPX, in contrast to placebo and naproxen (PLC+NPX) in SBP with signs of radiculopathy, and as a function of gender. We hypothesized that the combination LDP+NPX treatment would outperform PLC+NPX; PLC+NPX treatment would show minimal efficacy and be similar or even inferior relative to effects observed in an SBP cohort with low-risk for pain persistence receiving no-treatment. Our secondary hypothesis was that response to LDP+NPX treatment would be gender dependent.

A parallel aim was the proof of concept of innovative tools for conducting clinical trials: targeting treatment to appropriate groups using neuroimaging-based biomarkers, use of objective markers for treatment efficacy, and network analysis to profile personality dimensions. Specifically, taking advantage of machine learning techniques and recent longitudinal identification of neurobiological markers that predict pain persistence in SBP individuals with radiculopathy^3,4,8^, we targeted treatments to a specific subtype of SBP: those at high-risk of transitioning to CBP (high-risk-SBP). The other subtype, low-risk-SBP, constituted a no-treatment arm (NoTx) and were simply observed for a similar length as those receiving treatment. This strategy not only enables testing the validity of our prediction model, it demonstrates the concept of targeting treatment to appropriate SBP subtypes, thus sparing patients from unnecessary side effects of treatment. Additionally, contrasting outcome differences between NoTx and treatment groups, while accounting for risk differences, provides additional evidence for efficacy. We also sought to explore the influence of treatment on the personality profile of back pain individuals, by examining network properties of pain-related questionnaire measures. Given that pain reports are subjective assessments, we additionally tested whether brain activity properties could provide objective evidence regarding treatment efficacy.

## RESULTS

A total of 125 SBP participants were screened between Feb 9, 2015 and March 28, 2018. 72 were eligible and were stratified between high-risk and low-risk of pain persistence. For stratification, we used a Bayes Naïve model that employed two brain imaging derived parameters shown to independently predict pain persistence in SBP patients with radiculopathy: regional fractional anisotropy, a white matter property reflecting fiber density and myelination^8^, and the functional connectivity between the right anterior hippocampus with limbic and pain-related regions (see model development details in **Supplementary Note 3**). Participants that, according to the model, had less than 60% probability of recovering naturally were deemed high-risk-SBP, while the others were classified as low-risk-SBP.

Out of 72 eligible participants, 61 were identified as high-risk (n=61) and were one-to-one randomized between treatment groups: LDP+NPX (n=21 completed) and PLC+NPX (n=28 completed). **Fig. 1** describes the trial profile, subject drop-outs and distributions, across the three study arms. After three months of treatment, treatment was ceased but participants were followed for another three months in order to assess long-term effects. Those at low-risk (n=11) were not given any treatment (NoTx group, n=10 completed) and were simply observed for a similar amount of time (6 months) as those randomized to treatment. All subjects who completed the study (n=59) were included in primary analyses. Safety analyses included all participants who received at least one dose of study medication. We observe no notable imbalances in baseline characteristics (age, gender, ethnicity, body mass index, income, education, pain duration) between the three groups (**Supplementary Table 1**). Mean age of the overall population was 47 years, and 41% of participants were females.

**Figure 1.**
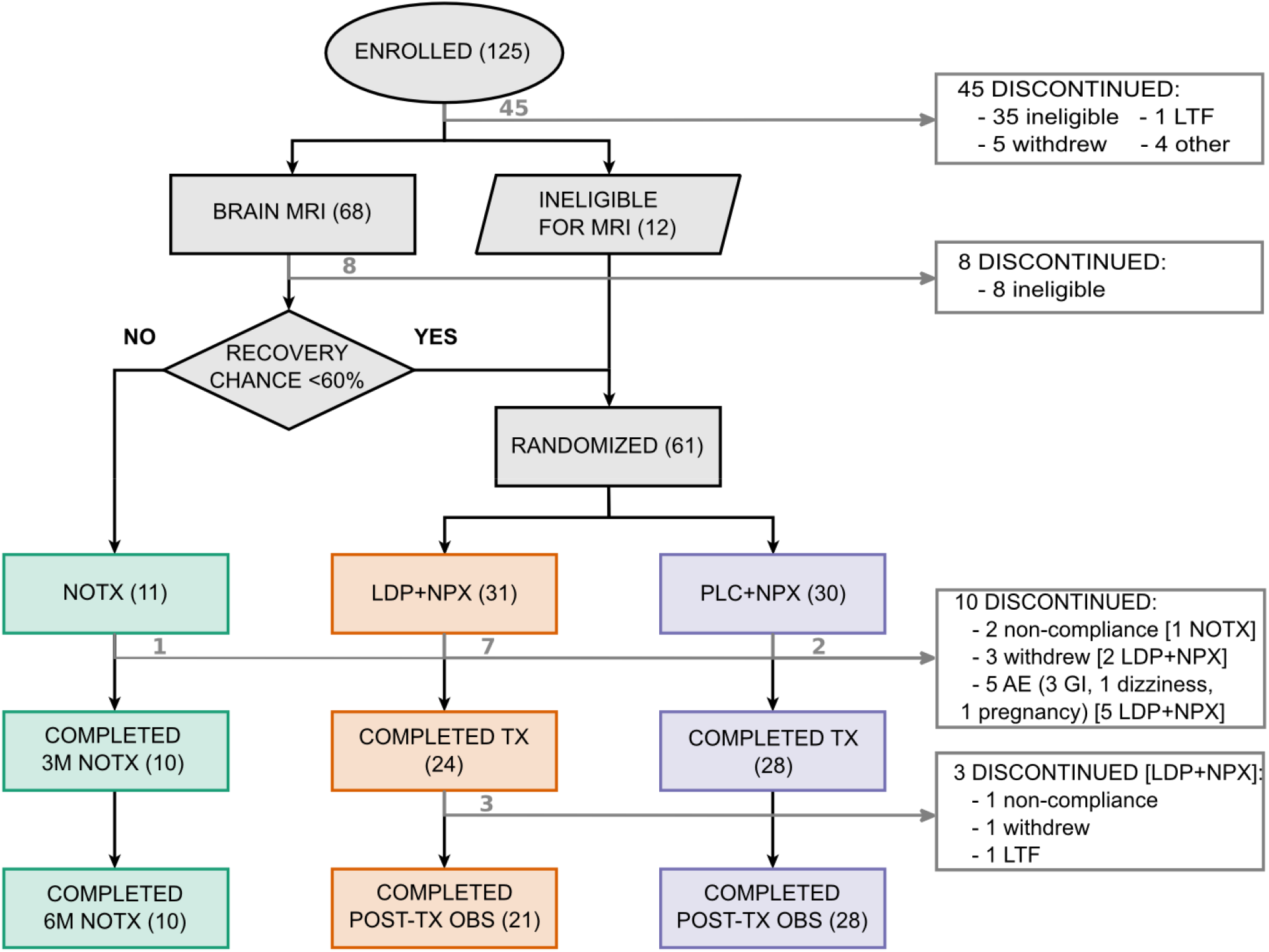
Trial profile. The diagram shows the numbers of subjects who met the criteria for inclusion in, or exclusion from, the study and their distribution among the three study arms. Entry in the treatment groups required a probability of recovery lower than 60%, according to a prediction model based on the baseline brain scans. LDP+NPX, levodopa/carbidopa + naproxen; PLC+NPX, placebo + naproxen; TX, treatment; NoTx, no-treatment; OBS, observation; AE, adverse event; GI, gastrointestinal distress; LTF, lost to follow.

### Both active treatment types result in sustained pain relief

Our main outcome measure was a *Numerical Rating Scale (NRS)*,^*19*^ where “0” corresponds to no pain and “10” indicates worst possible pain. On this scale, participants provided ratings of their current pain up to three times a day during study participation (6 months), via a smartphone app (phone NRS).

We computed treatment-related percentage residual pain relative to the average phone-NRS score from the week preceding start of treatment, for every subject and every day, and then averaged across subjects for each treatment type (**Fig. 2a**). Over the treatment period, we observe a rapid decrease in back pain intensity that, in the NoTx group, stabilizes (with large fluctuations) at around 80% residual pain, while in the LDP+NPX and PLC+NPX groups residual pain continues to decrease for the duration of treatment, reaching a residual pain of about 50%. A mixed-model of time course of pain ratings showed a significant time by group interaction (p<0.001), where NoTx was less effective than both active treatments. Remarkably in all three groups, the observed pain relief persisted after cessation of treatment, over the next 12-weeks.

**Figure 2.**
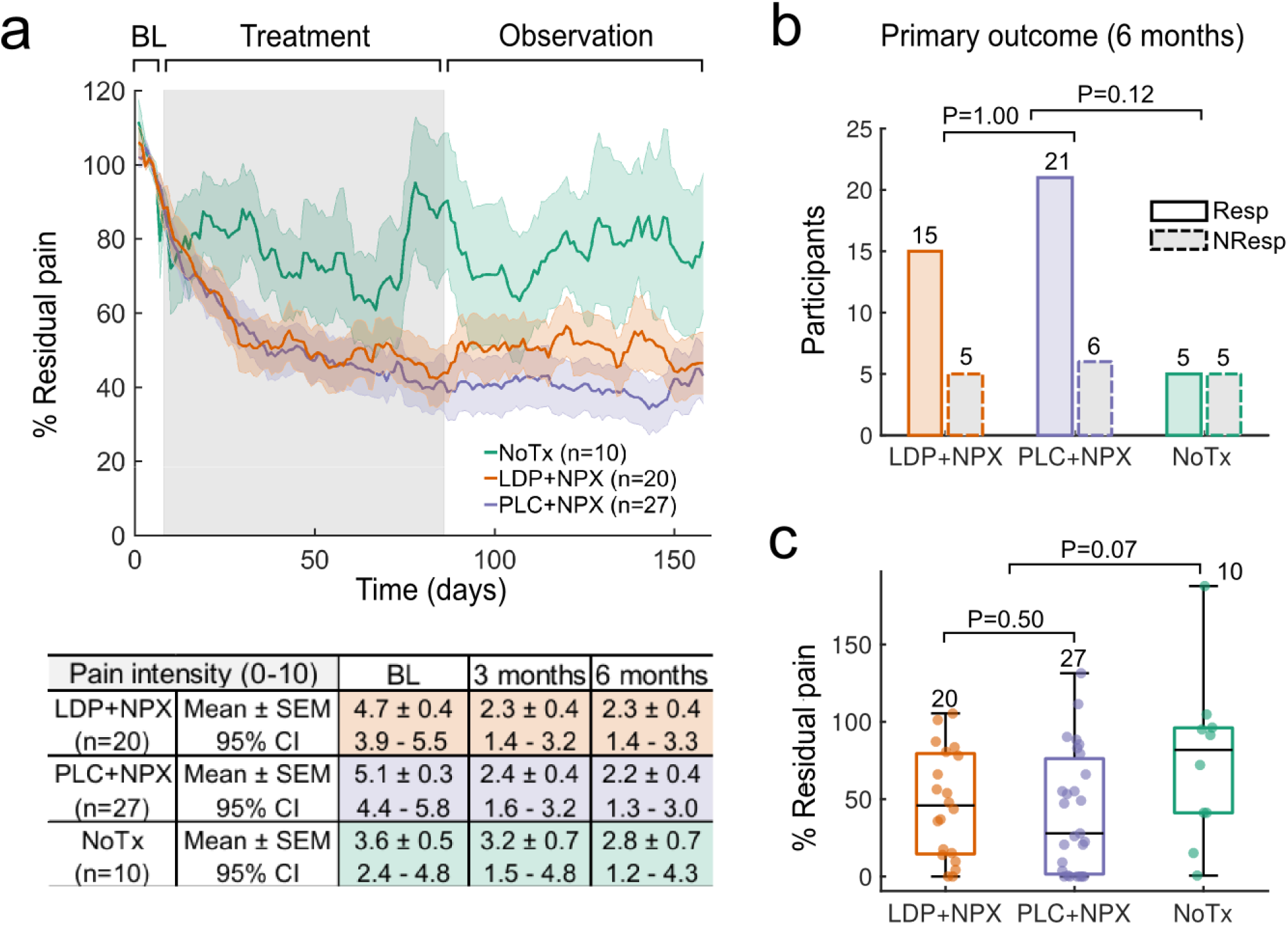
Residual pain trajectories and response rates. (**a**) The line plots depict daily average residual back pain intensity trajectories, per study arm: NoTx (green, n=10), LDP+NPX (orange, n=20), and PLC+NPX (purple, n=27) during baseline, treatment and post-treatment observation phases (displayed are 7, 78 and 73 days of ratings for each phase, respectively). There was an interaction between treatment and time (repeated measures mixed model ANOVA, F(314,8478)=1.54, P<0.001), with NoTx residual pain being greater than LDP+NPX (P=0.09) and greater than PLC+NPX (P=0.02, post hoc Dunnet test). Gre background indicates treatment-phase. Shaded areas represent +/- SEM. The table summarizes one-week group-averaged backpain (0-10 scale) at baseline and at three and six months, as a function of treatment type (mean and standard error, SEM, as well as 95% confidence intervals, CI, are shown). (**b**) Response rates in the LDP+NPX group (75% of subjects were responders) were not different from PLC+NPX (78% were responders) [Fisher’s exact test, P=1.00; with response criteria of 20% or greater reduction in mean pain intensity from baseline to six months, mean pain over one week of ratings]. A trend for superior response rates was seen in the treated population, relative to NoTx (Fisher’s exact test, P=0.12). (**c**) Residual pain at six months (based on one-week average of ratings) was not statistically different between study arms but showed a trend [Kruskal-Wallis test, P=0.15]. Comparing between NoTx and the two treatments together, treated individuals presented larger improvement in pain intensity (Mann-Whitney U test, P=0.07; medians, quartiles and ranges are shown). (At study closing and before data analyses, based on our original power calculation and total retention we declared P=0.1 as positive evidence for efficacy). Box plots indicate median, quartiles, and range, while numerals indicate number of subjects. BL, baseline; LDP+NPX, levodopa/carbidopa + naproxen; PLC+NPX, placebo + naproxen; NoTx, no-treatment; Resp, responders; NonResp, non-responders.

Contrary to our hypothesis, LDP+NPX did not yield superior pain relief relative to PLC+NPX at 6 months in high-risk individuals (**Fig. 2b**). However, both treatment arms (LDP+NPX and PLC+NPX) had about 75% of participants recovering (using an *a priori* defined criterion of ≥ 20% decrease in one-week average phone-NRS from baseline to 6-months; a clinically significant effect size for subacute and chronic back pain relief^20^), while the NoTx individuals, who were classified as low-risk, had a 50% recovery rate (based on the same *a priori* criterion). Model expected recovery rate was 60% for NoTx group (see **Supplementary Fig. 1**). Relative to NoTx, both treatments together were associated with larger improvement in back pain intensity (individual subject, and group, one-week average residual pain at 6-months, **Fig. 2c**). This same pattern is also seen in the table, where one-week average ratings of pain are shown for the three groups at three time points: baseline, 3- and 6-months, and confidence intervals (95% CI) are also reported. In both active treatment arms, but not in NoTx, 95% CI of one-week mean pain shows no overlap in range between baseline and 6-months.

Overall, the NoTx results closely match expected outcome (20% decrease in pain, in 50% of participants) based on our brain-imaging model prediction. Given recent meta-analyses of efficacy of nonsteroidal anti-inflammatories (NSAIDs) in acute and chronic back pain^21-24^ and given that active treatments were limited to participants where brain-imaging model categorized them at high-risk, we expected PLC+NPX to show less analgesic efficacy than NoTx. Instead, we observe both PLC+NPX and LDP+NPX showing pain relief at a magnitude not observed previously for any drug treatment for acute or chronic back pain^21,23,25^. Moreover, pain relief was sustained for 3 months after treatment cessation implying that we may be blocking transition to chronic pain.

### Treatment by gender interaction

As the brain MCL circuitry is sexually dimorphic^17,18^ and because we presume that transition to chronic pain is critically dependent on properties of MCL and its interaction with the nociceptive drive following inciting events^26^, we hypothesized, a priori, that active treatment efficacy should be gender dependent. To this end, we sub-grouped active treatment pain ratings by gender, and contrasted outcomes testing for a treatment by gender interaction.

Averaged daily residual pain time course (**Fig. 3a**) showed a treatment interaction with gender (p<0.001). When residual pain was examined at 6 months (average of one-week, relative to one-week baseline pain), as a function of gender, we again observe gender by treatment interaction (P=0.007, **Fig. 3b**). In females LDP+NPX treatment was superior to males (p<0.001), and superior to PLC+NPX-treated females (P=0.06). Next, we explored number of responders to active treatment, at 3-months and 6-months, when response criterion was varied from 80% - 20% residual pain from baseline (**Fig. 3c**). All females receiving LDP+NPX were responders at six months, even when response criterion was set as high as ≥80% decrease in pain from baseline (residual pain ≤20%), while PLC+NPX was less effective and similar among males and females at all four response criteria. Remarkably, over four response criteria, we observe essentially no change in recovery rates between 3 and 6 months, indicating persistence of pain relief, across treatments and genders.

**Figure 3.**
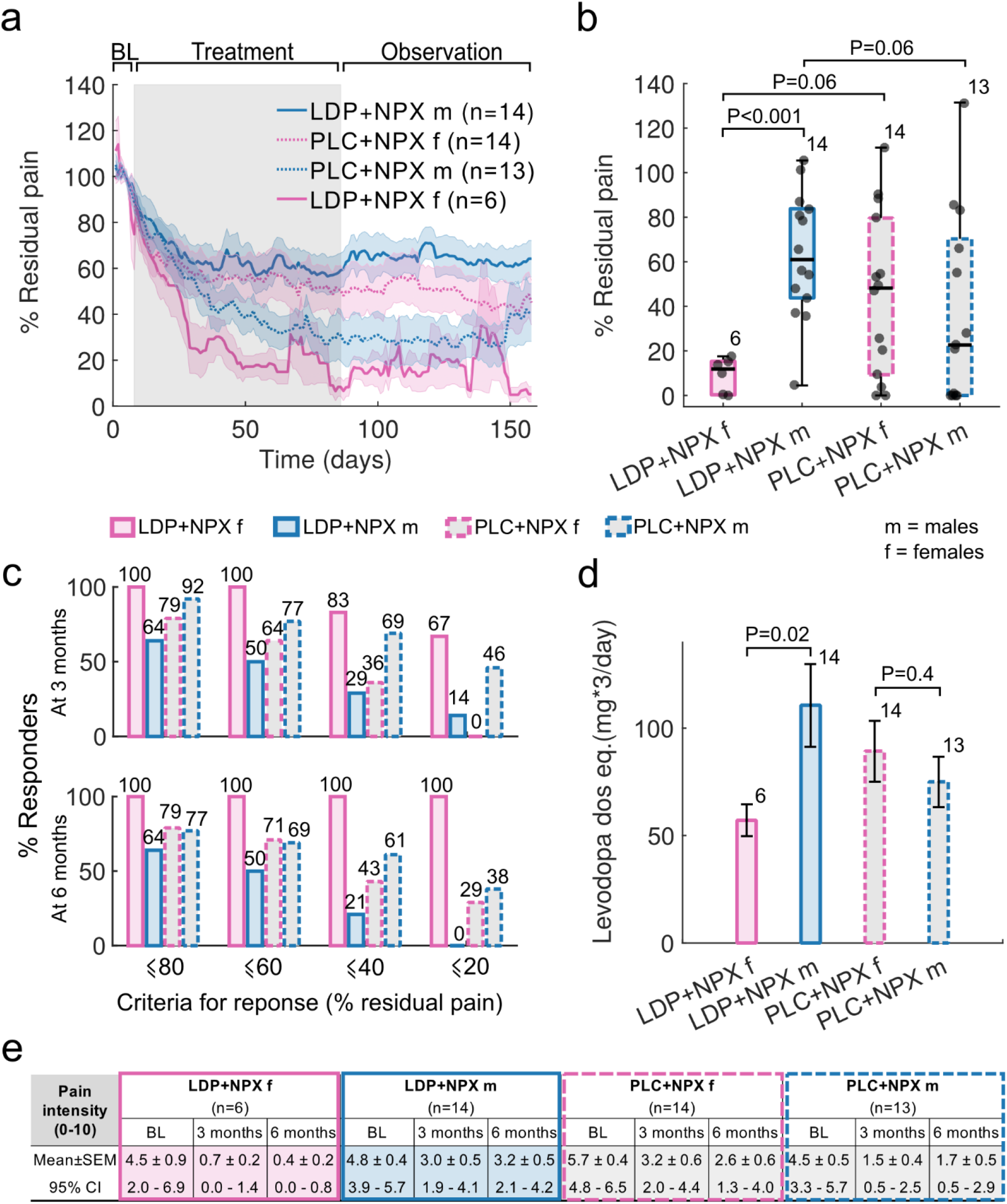
Residual pain trajectories and response rates, segregated by gender. (**a**) The line plot depicts average residual daily back pain intensity trajectories for females and males separately, per treatment arm. There was an interaction between time, treatment and gender (F(157,6751)=3.105, P<0.001, repeated measures mixed model ANOVA), with LDP+NPX females presenting lower residual pain than LDP+NPX males (P<0.05), and a tendency against PLC+NPX females (P=0.10), while PLC+NPX males had less residual pain than LDP+NPX males (P=0.06), according to post-hoc Tukey test. Shaded areas represent +/- SEM. (**b**) At six months, residual pain (one-week averaged pain ratings relative to baseline) showed a gender by treatment interaction (Two-way ANOVA, F(1,43)=8.16, P=0.007). In the LDP+NPX arm, females presented less residual pain than males at six months (post-hoc Mann-Whitney U test, P<0.001), indicating that LDP+NPX is more efficacious in females. Less residual pain was seen comparing these females against females in the PLC+NPX arm (post hoc Mann-Whitney U test, P=0.06). Contrasting males between treatments, PLC+NPX was more effective than LDP+NPX in males (post hoc Mann-Whitney U test, P=0.06). Box plots show median, quartiles, and ranges; while numerals indicate number of subjects per group. (**c**) Response rates at both three and six months across different criteria. LDP+NPX showed complete recovery across all thresholds in all females at 6 months, while PLC+NPX was less effective yielding similar response rates in males and females. (**d**) In LDP+NPX treated individuals, the average maximum administered dose (given three times/day) was lower in females than in males (two-tailed t-test, P=0.02). Levodopa dose equivalence was calculated according to number of placebo tablets. (**e**) The table summarizes group-averaged back pain at baseline and at three and six months (0-10 scale), as a function of treatment type and gender (mean and standard error, SEM, as well as 95% confidence intervals, CI are shown). LDP+NPX, levodopa/carbidopa + naproxen; PLC+NPX, placebo + naproxen; TX, treatment; NoTx, no-treatment; SEM, standard error of the mean.

Given the inexistence of previous evidence regarding efficacy of LDP+NPX in human pain management, appropriate dose for this combination treatment was unknown. Hence the current trial had a dose escalation design, which ranged from 12.5mg/50mg three times/day to 50mg/200mg carbidopa/levodopa, depending on the observed effects of treatment. Dose titrations were done in a blinded fashion. This design enabled calculating the LDP+NPX dose, or equivalent PLC+NPX dose, as a function of gender (**Fig. 3d**). We observe that, on average, females treated with LDP+NPX were given half of the carbidopa/levodopa dose received by males (P=0.02), despite no gender differences in mean body mass index.

When we examine mean back pain intensity and its range, as a function of gender and treatment type, we again observe that the largest decrease in back pain is seen in females treated with LDP+NPX (**Fig. 3e**).

In males, PLC+NPX seems to be more efficient, while LDP seems to interfere with the effect of NPX (**Fig. 3a, b, c, e**). Baseline pain duration, pain intensity, spread of pain on the body, and pain-related questionnaires showed no influence on these outcomes (**Supplementary Fig. 4** and **Supplementary Table 5**).

Overall, we find a consistent pattern of LDP+NPX being more effective in females using half the dose of males, and PLC+NPX being more effective in males. Still, in all four sub-groups (gender and treatment type) the observed decrease in pain during treatment persisted for the next three months of observation. The latter again reinforces the notion that we seem to be blocking transition to chronic pain.

### Change in back pain personality with treatment

In addition to daily phone ratings, during each visit, participants pain characteristics were assessed by rating their current pain intensity (NRS), and by questionnaires: 1) *Pain Sensitivity Questionnaire;*^*27*^ 2) *Pain Disability Index;*^*28*^ 3) *PainDETECT;*^*29*^ 4) *McGill Pain Questionnaire – Short Form;*^*30*^ 5) *Pain Catastrophizing Scale*;^31^ 6) *Pain Anxiety Symptoms Scale;*^*32*^ 7) *Beck Depression Inventory;*^*33*^ 8) *Positive and Negative Affect Scale*.^*34*^ Rather than examining treatment effects on each of these measures and their subscales, we opted for dimensionality reduction techniques, using a clustering algorithm, and constructing a network that summarizes participants’ personality profile. We then examined treatment effects on this network. This analysis is considered secondary and explores the influence of treatment on global personality properties of back pain individuals.

Baseline back pain personality network was derived from all participants’ measures correlation matrix (n = 116) (**Fig. 4a**). Five communities were identified, and were labeled based on member scales: 1) pain intensity; 2) pain sensitivity; 3) pain quality; 4) pain psychology; and 5) negative affect. Following a threshold to only keep correlations with p<0.01, we observe that these communities were tightly linked with each other at baseline (**Fig. 4b**). At six months, the network structure was disrupted, depending on treatment type (**Fig. 4c**). Treatment with LDP+NPX decreased correlation strengths throughout the network both in females and males (**Fig. 4d, f**), was associated with increased network modularity (**Fig. 4e**), and dissociated intensity measures from affective and pain-psychological factors (**Fig. 4c**).

**Figure 4.**
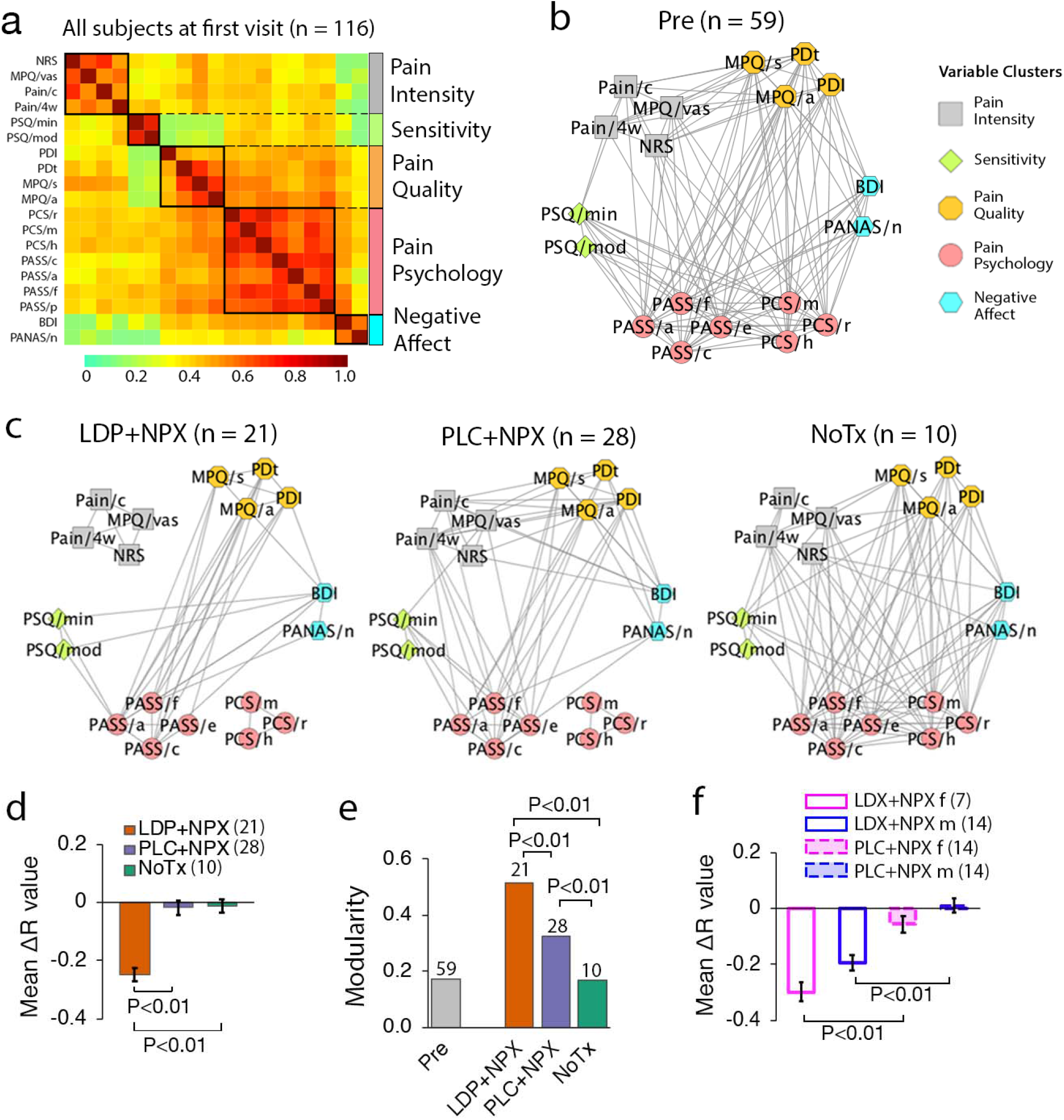
Treatment effects on backpain personality properties. (**a**) Correlation matrix for 19 questionnaire subscale measures. Clustering analysis identified 5 communities, based on data from 116 subjects (all participants available) at the first visit. Communities were labeled according to their component measures: pain intensity, pain sensitivity, pain quality, pain psychology, and negative affect. (**b**) Pre-treatment network graph displays interrelations between the five communities, highlighting within- and inter-cluster associations. Edges represent correlations with p<0.01. (**c**) At six months the network structure was disrupted in treated groups, whilst remaining intact in the NoTx group. Specifically, pain intensity and catastrophizing communities dissociated from the network in the LDP+NPX group. (**d**) LDP+NPX treatment decreased the strength of correlations (ΔR) between measures, relative to PLC+NPX and NoTx groups. (**e**) Network modularity increased in the treatment groups at 6 months, and was largest in LPD+NPX. (**f**) Relative to PLC+NPX, LDP+NPX decreased the strength of network correlations in both males and females. In **d** and **f**, error measures were based on permutation testing (10,000 repeated random resampling); error bars are SEMs. LDP+NPX, levodopa/carbidopa + naproxen; PLC+NPX, placebo + naproxen; NoTx, no-treatment. NRS, numerical rating scale; MPQ, McGill Pain Questionnaire - Short Form: visual analog scale (MPQ/vas), sensory (MPQ/s) and affective components of pain (MPQ/a); Pain/c, current pain (PainDETECT); Pain/4w (PainDETECT), average pain over the past four weeks; PSQ, Pain Sensitivity Questionnaire: minor (PSQ/min) and moderate (PSQ/mod); PDI, Pain Disability Index; PDt, PainDETECT; PCS, Pain Catastrophizing Scale: rumination (PCS/r), magnification (PCS/m), and helplessness (PCS/h); PASS, Pain Anxiety Symptoms Scale: cognitive suffering (PASS/c), escape-avoidance behaviors (PASS/e), fear of pain (PASS/f), and physiological symptoms of anxiety (PASS/a); BDI, Beck Depression Inventory; PANAS, Positive and Negative Affect Scale: negative (PANAS/n).

The back pain personality network was thus modified most with LDP+NPX treatment, and in females treated with LDP+NPX, fractionating the pain intensity community from the rest of the network.

### Objective brain functional correlates for treatment by gender interaction and for pain relief

Seeking to test objective neurobiological correlates to the subjective reports of treatment effects we used functional magnetic resonance images (fMRI), which were collected in most participants at baseline and six months later, to extract relevant brain properties. Candidate markers of treatment efficacy and gender dimorphism were predefined as the extent of information sharing, at rest, between three key regions of interest: the right nucleus accumbens (NAc), medial prefrontal cortex (mPFC) and anterior insula (Ins). Specifically, Ins functional connections with mPFC and NAc (Ins-mPFC, Ins-NAc) were hypothesized to reflect pain intensity, as reported in multiple previous studies,^8,35,36^ while mesolimbic connectivity (mPFC-NAc) was hypothesized to reflect gender specificity.^17,18^ Thus, for each participant we computed the connectivity strength between each pair of seeds, using previously reported region coordinates^3,37^, yielding three measures (correlation strengths between Ins, mPFC, and NAc activity at rest) from fMRI scans collected before and after treatment, and related these to our primary outcomes. No additional brain analyses were performed.

We found that these connections reflected distinct aspects related to treatment efficacy (**Fig. 5**). NAc-mPFC functional connectivity presented gender dependency at baseline (P=0.03), and in response to treatment it showed an interaction between gender and treatment (P=0.05, **Fig. 5a**). Across both active treatments, Ins-NAc (P=0.05) and Ins-mPFC (trend, P=0.16) functional connectivity changes over six months were associated with residual pain at six months (**Fig. 5b, c**). Therefore, predefined brain network properties yielded unbiased and objective correlates for treatment efficacy and gender dependence.

**Figure 5.**
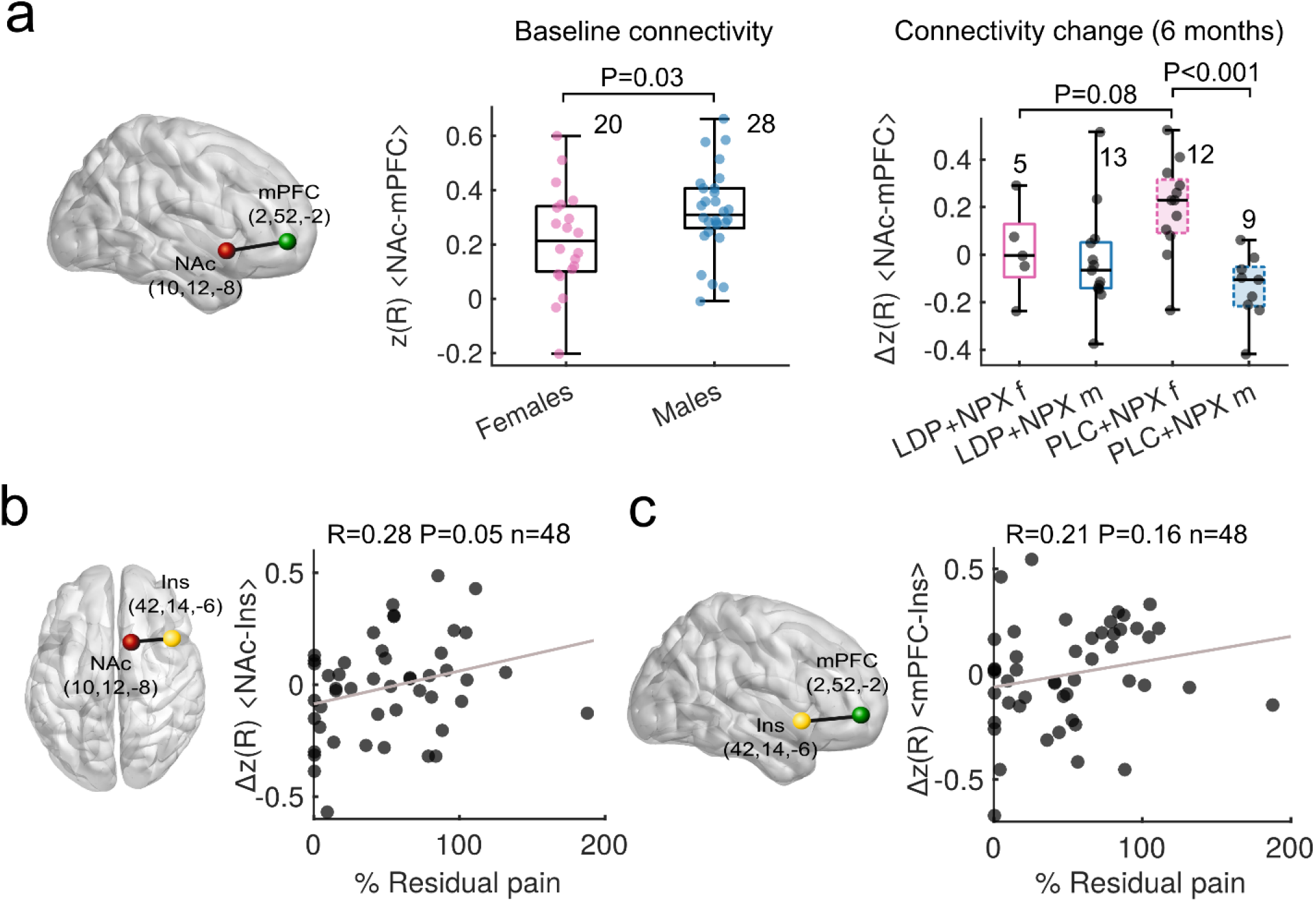
Resting state connectivity of three pre-defined regions in relation to pain, gender and treatment. (**a**) Functional connectivity strength between NAc and mPFC (NAc-mPFC; z-transformed correlation strength, Δz(R)) was weaker in females at baseline (Mann-Whitney U test, P=0.03). At six months, there was an interaction between gender and treatment on the extent of change in NAc-mPFC connectivity strength (Two-way ANOVA, F(1,35)=5.04, P=0.031). NAc-mPFC connectivity change was different between genders in the PLC+NPX group (post hoc Mann-Whitney U test, P<0.001) and between females in each treatment arm (post hoc Mann-Whitney U test, P=0.08). (**b**) Change in NAc-Ins functional connectivity strength was positively correlated with residual back pain at six months (Pearson R=0.28, P=0.05). (**c**) Change in mPFC-Ins connectivity presented a trend for association with residual pain, after regressing out contribution from age (Pearson R=0.21, P=0.16). LDP+NPX, levodopa/carbidopa + naproxen; PLC+NPX, placebo + naproxen.

### Relationship between brain-derived risk of chronic pain and treatment outcomes

Our brain anatomy and functional connectivity-based model predicting risk for chronic pain, calculated at time of entry into the study, successfully estimated residual pain six months later in the NoTx arm, based on the main outcome measure (phone NRS, P=0.05), as well as across other pain intensity measures (**Fig. 6, top row**). These associations were clearly disrupted in both LDP+NPX and PLC+NPX treatment arms, indicating that both treatments successfully disrupted the predicted natural trajectory of pain (**Fig. 6, mid and bottom rows**). Given that our predictive model was validated in the NoTx group, we can approximate expected recovery rate in the treated subjects from the model. The model approximates that only less than 25% of the high-risk group (treated with LDP+NPX or PLC+NPX) should have been recovering, while both of these treatments resulted in ∼75% recovering. Thus, these results not only validate our classifier but they also provide additional confidence for treatment efficacy.

**Figure 6.**
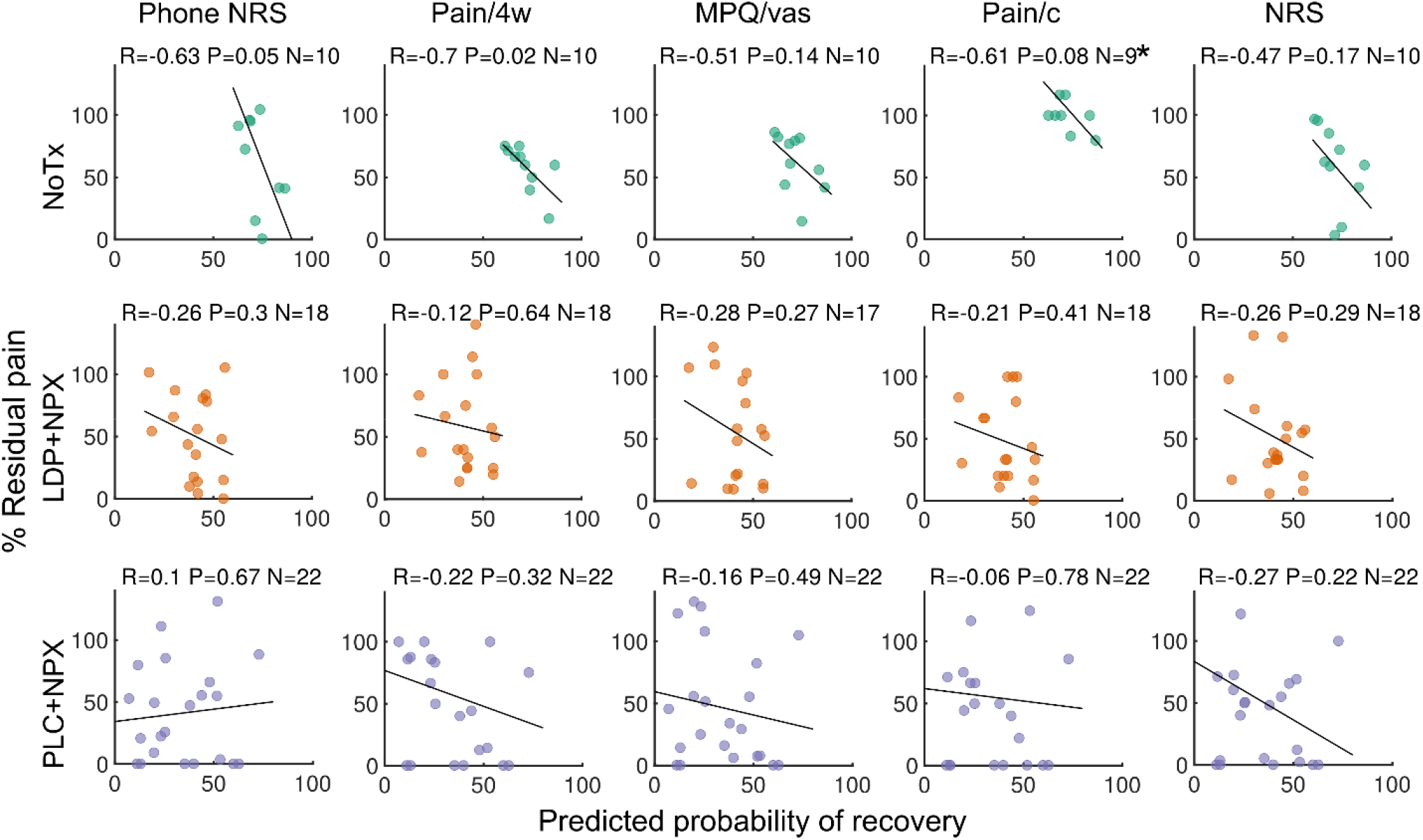
Model-based predicted recovery is disrupted by treatment. Scatter plots depict model-based predicted probability of recovery versus residual back pain at 6 months, for each treatment type, and for 5 measures of back pain intensity. There is a consistent negative correlation between predicted probabilit of recovery and residual pain in the NoTx group, across pain intensity measures, indicating that the predictive model used to stratify patients *a priori* performed well (top panel). Such associations were absent in both LDP+NPX and PLC+NPX treatment groups (lower two panels), suggesting that treatment successfully disrupted what would have been the natural pain trajectories for these participants. *There was one Pain/c outlier (with residual pain ∼600%) in the NoTx group who was removed; Pearson R is displayed. LDP+NPX, levodopa/carbidopa + naproxen; PLC+NPX, placebo + naproxen; NoTx, no-treatment; NRS, numeric rating scale; MPQ/vas, visual analogue scale of McGill questionnaire; Pain/c, current pain (PainDETECT); Pain/4w, average pain over the past four weeks (PainDETECT).

### Adverse events

As the study is the first clinical trial of the combination therapy, documenting adverse events was one of its primary aims. Clinical adverse events were reported by 19 (61.3%) and 17 (56.7%) participants in the LDP+NPX and PLC+NPX groups, respectively, with no statistically significant differences between treatments. In both treatment arms multiple adverse events were consistent with those commonly observed for non-steroidal anti-inflammatories^43^. Three LDP+NPX participants reported serious adverse events, all of which were judged to be unrelated to the medications used (see **Supplementary Tables 8-10**).

## DISCUSSION

In this proof of concept neuroimaging-based trial involving SBP patients, we demonstrate novel concepts regarding transition to chronic pain: 1) a 12-week treatment resulted in pain relief sustained for the next 12 weeks, suggesting blockade of transition to chronic pain; 2) LDP+NPX treatment seems safe, highly effective in females and fractionated pain intensity components from the back pain personality network; 3) brain functional connectivity reflected gender dimorphism and therapy correlates, providing complimentary objective measures for efficacy; 4) risk for chronic pain measured from brain parameters at time of entry into the study predicted back pain six months later in NoTx, but not in the treated subjects; thus establishing that such models can be tools towards individualizing treatments, in addition to providing additional independent and objective evidence regarding treatment efficacy.

Our trial was designed with the notion that SBP with radiculopathy constitutes a highly vulnerable population for developing CBP, who are within a critical time window from pain onset, when associated central reorganizations may be reversible. Therefore, participants were treated for a long-duration and monitored for long-term persistence of efficacy. The results confirm this concept, based on daily ratings of back pain, profiling of SBP personality, and through objective brain biomarkers and brain-derived model for risk of chronic pain. Thus, the approach seems to reset the back pain into a new and persistently improved level for the next three months, suggesting that the extent of reorganization accompanying treatment should persevere in the longer term.

Although the number of subjects studied remains small and, on the aggregate LDP+NPX was not superior to PLC+NPX, in SBP females, this treatment seems more efficacious than any therapies currently available for acute, subacute, or chronic BP^38,39^. At present, there are no sex-specific treatments for pain. Instead studies tend to either only include males or pool both sexes together for analyses of treatment outcomes^40^. Short-term NSAIDs are currently recommended for acute, subacute (first line), and chronic BP (first line)^41^, yet effects on pain intensity relative to placebo are small: −8.4% (95%CI −12.7 to −4.1) for acute BP, −3.3% (95%CI −5.3 to −1.3) for persistent BP, (weighted mean differences in pain reduction, with 95% confidence intervals, for 0-100 point scale at short-term follow-up)^39,41^. Relative to baseline, CBP patients show mean reduction of −14.3% (95%CI −16.0 to −12.6) following NSAID treatment^21^. Antidepressants and opioids are also used to treat persistent BP; however, these are not more effective than NSAIDs^21,25,42^. For non-specific back pain, meta-analysis show that the analgesic effect across different types of treatments are small, with 47% of patients having <10 points effect (on 0-100 point scale), and only 15% having point estimates of >20 points relief ^23^. In contrast, here we observe 50% of NoTx achieving >20% relief, and 75% of LDP+NPX or PLC+NPX groups achieving >20% relief. All SBP females in the present study had at least 80% improvement in their pain intensity after taking LDP+NPX, while males had better outcomes taking PLC+NPX (38% of males showing at least 80% improvement with PLC+NPX, while no males taking LDP+NPX had 80% relief). The large difference in efficacy seen between female and male SBP is consistent with the literature regarding MCL being sexually dimorphic,^17,18^ which in turn is backed by our female participants having lower resting state NAc-mPFC connectivity strength at entry into the study. Thus, this is the first evidence suggesting gender specific, effective treatment for back pain, with exceptional outcomes for females.

In exploratory analysis, here we introduce the use of network analysis to profile personality dimensions of back pain patients. We observe that fundamental characteristics of the network topology are fractionated to different extents with each treatment type, and with gender dependence. This analysis provides a global overview of the pain personality of the subjects studied and shows that treatment modulates fundamental properties of the network, with much less effect on specific questionnaire unitary outcomes.

As LDP+NPX has not been previously studied in the context of human pain, it was important to carefully document associated adverse events. We observe no adverse effects specifically linked to LDP use. Instead, we only observe adverse events commonly related to NSAIDs. Current American and European guidelines for management of CBP recommend limited time of use NSAIDs due to adverse effects:^21^ shortest duration possible and up to three months, respectively. In meta-analysis mean duration for treating BP with NSAIDs was 5-7 days.^22^ Here we treated participants for three months and found that both PLC+NPX and LDP+NPX had unperturbed vital signs and acceptable adverse-event profiles, which were similar to those reported for NSAIDs in general,^43^ with the additional LDP showing minimal or no evidence of increased side effects. While full toxicity evaluation and long term effects remain to be tested in larger sample sizes, obtained results indicate that 12-week standard analgesic dose of NSAID use in males, and standard dose of NSAID plus pediatric doses of levodopa/carbidopa in females have a high likelihood of blocking transition to chronic pain, at least for SBP with back pain of duration <20 weeks, and decrease intensity of back pain by about 50%. Importantly we see no evidence of the back pain duration moderating treatment outcome, thus it is also possible that such treatments may be efficacious in patients with longer durations of persistence of back pain, implying plausible efficacy even in CBP. It remains unclear whether these treatments also apply to other sub-acute pain conditions, especially other musculoskeletal pains.

The study does have important weaknesses that need highlighting. Foremost the results remain based on a small number of participants, especially SBP women treated with LDP+NPX. As treatment efficacy was highest in the latter group, larger study of this effect is urgently needed. Given that the PLC+NPX showed much larger efficacy than initially hypothesized, it could no longer be treated as an adequate control for the LPD+NPX treatment. Therefore, future studies must also include additional controls, for example: a PLC-only arm; and short-duration (1-2 weeks) PLC+NPX and/or LDP+NPX controls to actually establish that the 3-month treatment period in SBP is a critical factors for blocking transition to CBP. A recent case report^53^, shows that in 2 females with back pain and restless leg syndrome low dose LDP resulted in relief from back pain. Thus, it is also important that future studies compare efficacy of LDP+NPX in contrast to LDP, especially in women.

Collectively, results presented here establish proof of concept that early long-duration treatment of back pain with naproxen and its combination with levodopa/carbidopa are viable treatments, with persistent efficacy of at least three months after treatment cessation. It should be noted that dense monitoring of pain (daily ratings over ∼6 months) combined with a blinded trial and with objective brain correlates of primary outcomes, render the results highly unlikely to be due to a consequence of random effects. Additionally, the approach provides novel concepts regarding conducting clinical trials especially for subjective conditions, like pain. Still, these findings need to be replicated in a larger population before changing clinical practice.

## Data Availability

Data will be available post publication.

## Contributors

DR wrote the manuscript, lead the conduct of the trial, performed data analyses and data collection. AVA wrote the manuscript, originated the idea, lead on study design, supervised conduct of the trial and analyses. TJS lead on study design and conduct of the trial, evaluated toxicities and provided clinical oversight. MNB supervised and participated in data analyses. PT and MG lead the conduct of the trial and performed data collection. LH and KW participated in data analyses. BP collected data and participated in data analysis. EVP collected data and participated in analytic design. TA, RJ, SB, AB collected data and participated in the conduct of the trial. JWG was the trial statistician.

## Acknowledgements

We are thankful to all Apkarian lab members who contributed to this study with their time and resources. We would also like to thank all individuals that participated in this research. The study was funded by National Institute of Dental and Craniofacial Research (Blue Print grant number R01DE022746) and in part by National Institute on Drug Abuse (P50 DA044121).

## Data sharing

Data from our previous studies are already available on OpenPain (www.openpain.org). Data from the present study will be presented in more than one manuscript and will eventually be made available on OpenPain, upon completion of these manuscripts. The full study protocol may be made available upon request.

## METHODS

This 24-week double-blind parallel randomized controlled trial was conducted at Northwestern University (Chicago, USA). Protocol and informed consent form were approved by Northwestern University IRB as well as NIDCR/NIH. All enrolled participants provided written informed consent. Safety and trial oversight were monitored by an independent data safety monitoring board and a clinical research organization, with NIDCR/NIH oversight. The trial was registered on ClinicalTrials.gov, under registry NCT01951105.

### Participants

Individuals with a recent onset of low back pain, SBP, were recruited through online social media, local advertising, and via Northwestern Medicine Enterprise Data Warehouse. Criteria for enrollment included: history of low back pain with duration between 4-20 weeks, with signs and symptoms of radiculopathy, average reported pain intensity greater than 4 (on an NRS scale from 0 to 10) on the week before baseline assessments and the week preceding treatment start. We should note, however, that n = 18 subjects fell short of the back pain intensity criterion. Subjects were excluded if they reported other chronic painful conditions, systemic disease, history of head injury, psychiatric diseases or more than mild depression (score >19, according to Beck’s Depression Inventory^33^). See **Supplementary Note 2** for a full description of the inclusion and exclusion criteria.

### Trial Design

Eligible subjects first underwent a brain imaging session and, using predefined brain-derived parameters, were stratified into high and low-risk subgroups for transitioning to CBP. Those in the low-risk category, who had 60% or higher probability of recovering naturally, entered the NoTx arm; the rest, who had less than 60% probability of recovering naturally, were randomized between a placebo-control arm, and a pharmacological treatment arm. A flexible dose escalation procedure was used and both researchers and participants were blinded to treatment type. Study medications were tapered off at week 12, and participants continued to be evaluated for another 12 weeks. At final visit, participants underwent a second brain scan.

The design was setup to evaluate safety, efficacy and gender dependence of the combination of carbidopa/levodopa (12.5mg/50mg –25mg/100mg – 50mg/200mg, dose escalation based on response) plus naproxen 250mg (LDP+NPX) administered three times a day, compared to placebo plus naproxen 250mg (PLC+NPX) administered three times a day.

### Modeling risk for chronic pain and stratifying participants

We used a Naïve Bayes classifier to estimate probability of recovery from back pain for each participant, before they entered the treatment phase. The classifier was trained using data from our previous longitudinal study in SBP and employed two brain markers to predict risk for CBP. These were: the mean fractional anisotropy (FA) of white matter regions of interest that together predicted persistence of low-back pain at one year in previous work,^8^ and the number of functional connectivity links (degree count, De) between the right anterior hippocampus, and limbic and pain-related areas (**Supplementary Note 3**). Every participant eligible for MRI underwent an initial brain scan, and brain derived FA and De measures were entered into the equation above to identify probability for recovery. This measure was used to stratify participants into treatment and no-treatment arms. Participants who could not be scanned (n=12) were assumed to belong to the persisting category (as expected incidence rate for persisting category was ∼80%) and were entered into the treatment arm. Further model calculation details are described in **Supplemental Note 3**.

### Randomization and masking

Eligible individuals were assigned to treatment arms (LDP+NPX, PLC+NPX) based on a computer-generated permuted block randomization scheme, with block size randomly varying and an allocation ratio of 1:1. Allocation concealment was ensured by utilization of sequentially numbered containers. An unblinded individual from Northwestern University Clinical and Translational Sciences (NUCATS), with no other role in the study, was responsible for assuring proper medication assignment to each container. The randomization code was maintained by NUCATS and was available in cases of emergency or clinical situations in which knowing the treatment allocation would make a difference in the safety or management of a subject. In such a circumstance, the allocation assignment was made available after consultation with the site investigator and the principal investigator. This procedure was implemented for one participant who developed a serious adverse event during the treatment period. At study conclusion, after database lock, the randomization code was made available to researchers analyzing data. Participants received 2 capsules on a three times a day (TID) schedule and one capsule once a day (QD). TID medications included one capsule of naproxen and one capsule of either placebo or some dose of carbidopa/levodopa. Omeprazole 40g was taken QD in the morning, as a preventive measure against gastric adverse effects of naproxen. In order to assure that participants took their study medication as designed, the naproxen and omeprazole were placed in a separate colored capsule from the carbidopa/levodopa. Each colored capsule was dispensed in separate containers and participants were asked to take one capsule from each container. Acetaminophen was available as a rescue medication and all participants were given equal amounts.

### Procedures

The study consisted of eight visits spread over ∼28 weeks, which included an initial screening/monitoring period (∼2 weeks), followed by a treatment period (12 weeks) and a post-treatment observational period (12 weeks). Safety and adverse events were assessed at each clinical visit, including vital signs. At each visit, researchers reminded participants to take their medication according to instructions. Safety was also assessed through blood screenings at baseline, while blood pressure, pulse and temperature were assessed at all study visits.

### Dose escalation protocol

Treatment with carbidopa/levodopa was titrated up to 12.5mg/50mg three times/day over one week and then continued at that level for 4 weeks. If by the end of the initial 4-week period the participant “responded” [had >20% decrease in pain intensity from the average of all phone NRS ratings collected at baseline (between visit 1 and visit 3) to the average of all phone NRS ratings during those 5 weeks], the participant was maintained on that dose for the duration of the treatment period (12 weeks total). If there was no response, the carbidopa/levodopa dose was increased to 25mg/100mg three times/day for the following 4 weeks, at which time the pain status was re-evaluated (based on the average of all phone NRS rating during those four weeks, against baseline average). Again, if a response occurred, that dose was maintained in a blinded manner for the following 4 weeks of treatment; if not, further dose-titration carbidopa/levodopa occurred to 50mg/200mg three times/day for the final 4 weeks. When a participant experienced an adverse event at higher doses, the participant was given the next lower dose that s/he was able to tolerate and then maintained on that dose for the remainder of the 12-week dosing period. Naproxen dose (250mg three times/day) remained constant for all participants throughout the study, except it was not given during the tapering down at the end of the study.

### Monitoring pain intensity with phone app

The main outcome measure was a *Numerical Rating Scale (NRS)*,^*19*^ where “0” corresponds to no pain and “10” indicates worst possible pain. Participants were instructed to provide such ratings three times a day, for at least one week prior to randomization, and throughout study participation, via a smartphone app (phone NRS), which contained the following instructions: “Please rate your current level of pain”.

### Phone NRS preprocessing

Percentage residual pain at three and six months were computed based on the average phone-NRS score from the week preceding treatment and the final week of treatment or post-treatment. Thus, 100% residual pain reflected no change from baseline levels; while 0% residual pain reflected complete recovery. Participants were deemed responders if residual pain at six months was 80% or less (representing 20% pain relief), as defined *a priori per protocol*.

Specifically, when an individual had more than three ratings in a given day, we only kept three: the first, the last and one rating in between. Next, the average of these ratings was computed for each day. For missing daily ratings, the mean from nearest neighbors were used to replace the missing value. Two subjects who completed the study were not included in the phone NRS analyses due to non-compliance in providing phone ratings.

For each subject, percentage residual back pain (*% Residual pain*_*i*_) at a given day *i* was computed as:

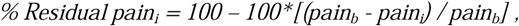

where *pain*_*b*_ is the baseline pain intensity, pre-defined as the average daily phone-NRS during the week preceding start of treatment, and *pain*_*i*_ is the mean phone-NRS at day *i*. Thus, 100% residual pain reflects no change from baseline levels; while 0% residual pain reflects complete recovery. Primary outcome measure was pre-defined as the average phone-NRS residual pain during the final week of the study (six months from start of treatment, *i*.*e*, three months after end of treatment). Participants were considered responders if they had 80% or less residual pain at six months.

We additionally examined outcomes at three months (that is, at end of treatment), which was defined as the average phone-NRS residual pain during the final week of treatment. For analyses of pain intensity trajectories, we included 7 days preceding treatment, 78 days during treatment and 73 days post-treatment.

### Other outcomes

In addition to phone ratings, during each visit, participants rated their current pain intensity (NRS), and were administered self-report questionnaires:

1. *Pain Sensitivity Questionnaire (PSQ)*,^*27*^ a 17-item instrument used to assess individual pain sensitivity - it is based on pain intensity ratings of hypothetical situations, which includes various modalities (heat, cold, pressure, pinprick) and measures (pain threshold, intensity ratings); It can be split into two subscales: one consisting of items referring to mildly painful situations (minor, PSQ/min), and one consisting of the items referring to moderately painful situations (moderate, PSQ/mod);
2. *Pain Disability Index (PDI)*,^*28*^ an assessment of physical impairment in relation to pain;
3. *PainDETECT*,^*29*^ *a 12-item assessment of neuropathic-like symptoms. PDt includes questions of current pain intensity (Pain/c) and subjective report of average pain intensity over the past 4-weeks (Pain/4w);*
4. *McGill Pain Questionnaire – Short Form(sf-MPQ)*,^*30*^ *a well-validated measure assessing both sensory and affective components of pain (MPQ/s and MPQ/a). It also includes a visual analog scale (VAS) of pain;*
5. *Pain Catastrophizing Scale (PCS)*,^31^ is a 5-point instrument to assess 13 thoughts or feelings on past pain experience. PCS yields three sub-scale scores assessing rumination (PCS/r), magnification (PCS/m), and helplessness (PCS/h);
6. *Pain Anxiety Symptoms Scale (PASS)*,^*32*^ measures fear and anxiety responses specific to pain. PASS consists of four aspects of pain-related anxiety: cognitive suffering (PASS/c), escape-avoidance behaviors (PASS/e), fear of pain (PASS/f), and physiological symptoms of anxiety (PASS/a);
7. *Beck Depression Inventory (BDI)*,^*33*^ is a 21-item instrument for measuring the severity of depression;
8. *Positive and Negative Affect Scale (PANAS)*,^*34*^ *has two mood scales, one measuring positive affect and the other measuring negative affect (PANAS/n). Each scale is rated on a 5-point, 10-item scale*.

These measures were considered secondary and were used to construct back pain personality profile and examine treatment effects globally.

We differentiated between adverse events (AEs) that occurred during treatment and those that occurred after treatment. However, to account for potential late onset AEs or those related to withdrawal of medication, we also considered AEs occurring during a certain window after treatment to be “during treatment”. Explicitly, we defined that AEs would be accounted as “occurring during the treatment period” if they were observed during the interval from the first dose of study drug to 28 days after the last dose of study drug, or end of study participation, whichever occurred first.

Information on gender, race and ethnicity were obtained using a standard National Institute of Health demographics form, which did not explicitly differentiate between gender and biological sex. Here we use the word gender, as that was the word used on the self-report form completed by participants.

Candidate brain markers of treatment efficacy and gender dimorphism were predefined as the extent of information sharing, at rest, between three key regions of interest: the right nucleus accumbens, medial prefrontal cortex and insula. Specifically, insula connectivity was hypothesized to reflect pain intensity, as found in previous study,^8^ while mesolimbic connectivity was hypothesized to reflect gender specificity.^17,18^ Thus, for each participant we computed the connectivity strength between each pair of seeds, yielding three measures from fMRI scans collected before and after treatment, and related these to our primary outcomes. No additional brain analyses were performed.

### Brain imaging acquisition

Data were acquired on a clinical 3T Siemens Magnetom Prisma whole body scanner equipped with a receive-only 64 channel head/neck coil. At both baseline and 6 months after randomization, participants underwent a high-resolution anatomical scan (T1-weighted MRI), one resting and one spontaneous pain rating functional MRI (fMRI), and a white matter fractional anisotropy (FA) assessment scan (Diffusion-weighted MRI). The entire MRI scan consisted of approximately 35 minutes of actual image acquisition, plus around 25 minutes for setting patients comfortably in the scanner, attempting to minimize their back pain, and for re-acquiring images in case of excessive motion.

#### T1-weighted MRI acquisition

High-resolution T1-weighted brain images were collected using integrated parallel imaging techniques (PAT; GRAPPA). The acquisition parameters were: voxel size: 1 × 1 × 1 mm^3^, TR = 2.3 s, TE = 2.40 ms, TI = 900 ms, flip angle = 9°, 176 sagittal slices and acceleration factor = 2. Phase encoding direction was anterior to posterior, and the duration of acquisition was ∼5 min.

#### Resting-state fMRI acquisition

Blood oxygen level-dependent (BOLD) T2*-weighted multiband accelerated echo-planar images were acquired at rest. Acquisition parameters were as follows: TR□=□555□ms, TE□=□22□ms, flip angle□=□47°, 64 slices acquired with interleaved ordering, FOV = 208 mm, matrix size = 96 × 104, spatial resolution: 2□×□2□×□2□mm^3^, acceleration factor: 8. Phase encoding direction was posterior to anterior. Slices were acquired with ascending order to preserve the continuity of connections. The acquisition lasted ∼10min, during which 1110 volumes were collected. Participants were instructed to keep their eyes open and to remain as still as possible during acquisition.

#### Spontaneous pain rating fMRI acquisition

Identical acquisition parameters and duration to resting-state fMRI were used to obtain BOLD T2*-weighted images while participants used a finger-spanning device to continuously rate and log the rate of their spontaneous back pain on a scale of 0–100, in the absence of external stimulation.^44^ Participants were instructed to keep their eyes open and to remain as still as possible during the scan.

#### Diffusion-weighted MRI acquisition

Multi-slice echo planar imaging with multiband excitation and multiple receivers was used to obtain diffusion-weighted images along 30 and 64 evenly spaced and non-collinear directions, with weighting factors of 700 s/mm^2^ and 2000 s/mm^2^, respectively. Two non-weighted volumes were acquired, each at the beginning of each scan. Voxel size = 2 × 2 x 2 mm^3^, TR = 3.5 s, TE = 92 ms, FOV = 230 mm, matrix size = 116, number of slices = 72, flip angle = 90°, multiband factor = 3, acquisition time ∼7 min.

### Brain imaging processing (for baseline model parameters)

For each subject, MRI data was processed within less than a week from the baseline acquisition and before the randomization visit, in order to extract brain parameters for patient stratification. The quality of each image modality was assessed for excessive motion and poor signal to noise ratio before preprocessing, using a robust quality control pipeline.^45^

#### Spontaneous pain rating fMRI: right hippocampus connections with limbic and “pain” regions

fMRI volumes were preprocessed using tools within the FMRIB Software Library 5.0.9 (FSL), and MATLAB R2016a. After removing the first 120 volumes of each spontaneous pain rating functional dataset for magnetic field stabilization, skull extraction using BET and slice-time correction were performed. fsl_motion_outliers was used to remove the effect of intermediate to large motion. Next, the remaining 990 volumes were filtered with a band-pass temporal filter (using Butterworth; 0.008 Hz < f < 0.1 Hz) and a non-linear spatial filter (using SUSAN; FWHM = 5 mm). Finally, nine vectors were regressed out, including the six parameters obtained from intra-modal motion correction using MCFLIRT, global signal (averaged over all voxels of the brain, over the 990 volumes), white matter signal (eroded white matter mask), and cerebrospinal fluid signal (eroded ventricular mask). These nine vectors were filtered with the Butterworth band-pass filter before being regressed from the time series to avoid recontamination.

The cleaned images were then linearly registered to the MNI template using FLIRT and down-sampled to 6×6×6 mm^3^ voxel size. A C-based, in-house program called “ABLM” (Apkarian Brain Linkage Map), previously described by Baria and colleagues,^46^ was used to compute the mean count of functional connectivity links (degree) between voxels within a predefined mask within the right hippocampus and limbic and “pain”-related regions. Link density threshold for calculation of degree was set to the top 10% of connections.

#### Diffusion MRI: mean FA

Preprocessing of diffusion-weighted images was performed using eddy_current to correct for eddy current-induced distortions and subject movement. Using DTIFIT the diffusion tensor was estimated in each voxel by linear regression and FA maps were derived. Following this, each FA map was non-linearly registered to the FMRIB58_FA template. Next, we extracted the mean FA value of a group of voxels that was previously identified as having predictive value for pain chronification^8^ and used this value in our Naïve Bayes model for stratifying participants between high and low risk SBP (further details about the model are given below).

### Brain imaging processing (for post-hoc analyses)

For post-hoc analyses of brain function, we used our most recent preprocessing pipeline, which was optimized for longitudinal investigation. After removing the first 20 volumes (corresponding to 11s) of each functional dataset for magnetic field stabilization, data were subjected to skull extraction using BET, slice-time correction, motion correction with MCFLIRT, intensity normalization and high-pass temporal filtering (0.008 Hz). Motion censoring was next performed by detecting volumes with framewise displacement (a measure of how much the head changed position from one frame to the next) larger than 1 mm, DVARS (indexes the rate of change of BOLD signal across the entire brain at each frame of data) with z-score larger than 2.3, or BOLD signal z-score > 2.3, and removing their adjacent volumes (−5 −4 −3 −2 −1 0 1 2 3 4 5).^47^ Signals from three vectors were regressed out, including global signal (averaged signal over all voxels of the brain, over the 1090 volumes), white matter signal (extracted from an eroded white matter mask), and cerebrospinal fluid signal (extracted from an eroded ventricular mask). Finally, the cleaned time series were band-pass filtered (0.008-0.1 Hz) to keep the low-frequency fluctuations of interest.

Functional image registration was optimized for longitudinal analysis by utilizing a two-step approach that minimizes within-subject variability. First, for each subject functional images from scan 1 and scan 2 were registered to each other using forward and backward halfway linear transformations (FLIRT, 6 degrees of freedom). This process allows optimal within subject alignment, with minimal displacement artifacts for both scans.^48^ Second, the midway template was registered to MNI152 space using non-linear registration (FNIRT). In order to minimize warping effects, all non-linear registrations were constrained by affine registration as described by Smith et al.^49^ Final images were spatially smoothed with a Gaussian kernel (2 mm sigma). Registered data were visually inspected to ensure optimal alignment.

### Statistical considerations and analyses

The predefined primary outcome measure for efficacy, as a function of treatment type and relative to gender, was the percentage of participants recovering from back pain. Fisher’s exact test was used. Additionally, for analysis of pain intensity trajectories repeated-measures ANOVA was employed. Our secondary endpoint was to test validity of the model used for stratifying SBP, and also examine changes in brain connectivity with treatment. Exploratory analyses were conducted to examine treatment effects on pain-related questionnaire sub-scales, using dimensionality reduction methods and network analyses. Binary outcomes are reported based on absolute and relative descriptive statistics, consistent with CONSORT guidelines.^50^ All statistical tests were two-sided.

#### Power analysis for primary outcomes

Because our primary dependent variable was binary (recovery from back pain or not), and to have sufficient power to detect treatment effects, we conducted power analysis based on comparison of independent proportions. Our previous study in SBP shows that in high-risk SBP ∼90% persisted with back pain if treated with standard of care (mostly occasional use of anti-inflammatories).^3^ Thus, for power calculations, we assumed that 90% of participants that receive placebo and naproxen would have persisting pain at study end. Calculations in G*Power 3.1.9.2 indicated that we would detect significant differences with 80% power if the probability of pain persistence is reduced to 67% or smaller (Type I error rate assumed at .05, two-tailed, *n* = 50 per group). We assumed a two-tailed comparison, even though there was no *a priori* reason for the placebo plus naproxen to outperform the test drug. We therefore planned to enter 126 participants, accounting attrition and for the no-treatment group.

Our second aim was to determine if there is a gender by treatment interaction effect. Specifically, we wanted to test the influence of gender on response to active treatment with carbidopa/levodopa and naproxen. Assuming no gender effect on the placebo and naproxen treatment response rate, we are left with the active treatment arm. Calculations in G*Power 3.1.9.2 indicated that we would detect an odds ratio greater than 2.7 (2.5-3.0) with ≥ 80% power with an overall sample size of 50 (assuming n = ∼25 for each gender). We assumed a two-tailed comparison because there was no *a priori* reason for a given sex to respond better to the active drug. Again, as a supplemental predefined analysis strategy, we assumed to be able to strengthen the analyses, by removing variance due to nuisance covariates and using longitudinal modeling. Thus, total planned participants needed to increase by an additional 60 subjects, making the total planned recruitment 186. However, due to financial constraints we could only recruit 125 participants, and as a result we determined (prior to any data analysis) that the primary outcome would be considered positive if comparison outcome passed the less stringent criterion of p<0.1, rather than p<0.05.

#### Model testing

In order to test the validity of our classifier which stratified participants between high and low risk of pain persistence, we examined the actual responses of the NoTx group at 6 months from entry in the study against the predicted response from our model. To strengthen the validation, in addition to testing the actual response based on our primary outcome measure (Phone NRS), we also checked actual versus predicted recovery with other measures of pain intensity.

#### Identifying brain markers for treatment effects

Three regions of interest were defined *a-priori*: the right nucleus accumbens (NAc), medial prefrontal cortex (mPFC) and right anterior insula (aIns). These were based on previous studies showing that: 1. Connectivity strength between mPFC and aIns encode pain intensity across pain conditions,^37^ and 2. NAc-mPFC connectivity strength is causally associated with transition from subacute to chronic back pain. Additionally, the NAc has been previously shown to be sex-dimorphic,^17^ thus we reasoned that its connectivity may provide evidence for the gender specificity of treatment response seen for back pain intensity.

Mean time series representative of these regions were extracted from 10 mm radius spheres around previously reported MNI coordinates: NAc (10, 12, −8), mPFC (2, 52, −2) and aIns (42, 14, −6).^3,37^ The Pearson correlation coefficient (R) was computed, 3 numbers/brain, between BOLD signals from each pair of regions to represent their connectivity strength (extent of information sharing).

#### Questionnaire based exploratory endpoints

Given the large number of additional questionnaires used to assess participants’ treatment response, we used dimension reduction methods and network analysis methods to summarize these outcomes. These results are deemed exploratory in nature.

#### Missing questionnaire data

In case of missing within-questionnaire items, these were replaced with the average of the remaining within-questionnaire scores, provided that the number of unanswered questions was less than 30% of all items in each scale. If more than 30% of the items were blank, subjects were excluded from statistical tests relevant to the given scale.

#### Clustering analysis

To resolve multicollinearity, dimensionality-reduction techniques were applied to pain-related questionnaires at baseline (visit 1). Variable clustering under VARCLUS algorithm (SAS Institute Inc. 2017e) computed Pearson correlation-based similarity between all 19 included measures and assigned these measures to five clusters. VARCLUS is an iterative algorithm that calculates a determination coefficient (R^2^) between each variable and a cluster (own R^2^), of which the variable is a member, and R^2^ between each variable and the next most similar cluster (next R^2^). The (1 – R^2^) Ratio is defined as (1 – own R^2^) / (1 – next R^2^), where a ratio greater than 1 means that the next closest cluster is more similar than the current cluster.

#### Network analysis

Each of the N=19 pain-related questionnaire measures was used as a node. Based on this division, we constructed an undirected connectivity matrix ***B****=****R***^*NxN*^ representing all subjects who completed study participation at baseline. First, the Pearson correlation (R) coefficient was calculated for each of the possible pairs of nodes to generate a connectivity matrix. Next, this matrix was thresholded to only keep significant connections (P<0.01) and binarized yielding an undirected adjacency matrix. This process was repeated for each group (LDP+NPX, PLC+NPX and NoTx) separately at baseline, and at six months to investigate long-lasting treatment effects. For network visualization we used Cytoscape an open source software ^51^

To characterize the structure of these adjacency matrices we computed modularity using the Brain Connectivity Toolbox (BCT).^52^ A module can be defined as a set of nodes that are densely connected among themselves but sparsely connected to other parts of the network. Modularity quantifies how well-defined these densely connected sets of nodes are within the network. From the different modularity algorithms available, we chose to use the fast and accurate multi-iterative generalization of the Louvain method, provided within BCT. Using this technique, we obtained a single unitary value between 0 and 1 representing the modularity of each network, where values closer to 1 indicate highly structured systems and values closer to 0 represent random networks. In order to deal with potential modularity degeneracy, modularity was computed over 100 repetitions and the average of these iterations was used as the final modularity measure.

We further investigated global network structure by examining changes in connectivity on the non-binarized network from baseline to six months and averaging these changes over the entire network to obtain mean ΔR.

#### Group differences

Modularity and mean ΔR values were compared between groups using a permutation test. First, for each pair-wise measure, the difference between the two groups was calculated as the actual group difference. Second, we identified the lowest common number of subjects and resampled the combined pool of the two conditions into two new groups. The values of these two resampled groups were calculated next. This process was repeated 10,000 times to generate a null distribution of the mean difference between the groups. The p-value of the actual group difference was calculated as the chance probability from the mean in the null distribution.

#### Software

Analyses were done using MATLAB 2016a, JMP Pro version 13.2 (SAS Institute, Cary, NC) and SPSS version 25. Safety analyses included all participants who received at least one dose of the study drug.

